# The Estimated Time-Varying Reproduction Numbers during the Ongoing Epidemic of the Coronavirus Disease 2019 (COVID-19) in China

**DOI:** 10.1101/2020.04.11.20061838

**Authors:** Fu-Chang Hu, Fang-Yu Wen

## Abstract

**Background:** How could we anticipate the progression of the ongoing epidemic of the coronavirus disease 2019 (COVID-19) in China? As a measure of transmissibility, we aimed to estimate concurrently the time-varying reproduction number over time during the COVID-19 epidemic in China.

**Methods:** We extracted the epidemic data from the “Tracking the Epidemic” website of the Chinese Center for Disease Control and Prevention for the duration of January 19, 2020 and March 14, 2020. Then, we specified two plausible distributions of serial interval to apply the novel estimation method implemented in the incidence and EpiEstim packages to the data of daily new confirmed cases for robustly estimating the time-varying reproduction number in the R software.

**Results:** The epidemic curve of daily new confirmed cases in China peaked around February 4–6, 2020, and then declined gradually, except the very high peak on February 12, 2020 owing to the added clinically diagnosed cases of the Hubei Province. Under two specified plausible scenarios for the distribution of serial interval, both curves of the estimated time-varying reproduction numbers fell below 1.0 around February 17–18, 2020. Finally, the COVID-19 epidemic in China abated around March 7–8, 2020, indicating that the prompt and aggressive control measures of China were effective.

**Conclusion:** Seeing the estimated time-varying reproduction number going downhill speedily was more informative than looking for the drops in the daily number of new confirmed cases during an ongoing epidemic of infectious disease. We urged public health authorities and scientists to estimate time-varying reproduction numbers routinely during an epidemic of infectious diseases and to report them daily to the public until the end of the epidemic.

## Introduction

The coronavirus disease 2019 (COVID-19) is now known to be caused by the severe acute respiratory syndrome coronavirus 2 (SARS-CoV-2). The COVID-19 was first reported to the World Health Organization (WHO) from Wuhan City, Hubei Province, China on December 31, 2019. China locked down Wuhan on January 23, 2020. Then, the WHO officially declared the COVID-19 epidemic as a Public Health Emergency of International Concern on January 30, 2020. After drastic control measures had been implemented with a high cost to combat the COVID-19 epidemic, President Xi Jinping of China made his first visit to Wuhan on March 10, 2020 to show that China turned the corner on coronavirus. China announced no new locally transmitted cases on March 19, 2020, and then ended its lockdown of Wuhan on April 8, 2020. Nevertheless, the WHO described coronavirus as a pandemic on March 11, 2020 due to its rapid and wide spread over the world outside China. Traveling oversea has dramatically shortened the social distances between countries worldwide, and thus the coronavirus spreads out more quickly and widely than ever before. At the same time, public panic spreads out even more drastically through social and news media. The epidemic of such a novel infectious disease has caused big damages to human health, medical facilities, economy, and society in several countries. Once the epidemic is likely to occur, appropriate control measures according to the source-transmission-host theory, including the nonpharmaceutical interventions (NPI) and pharmaceutical interventions (PI), should be implemented timely to lower such damages.^1^ In China, prompt and aggressive actions had been taken since the very early phase of the epidemic. In the other counties, areas, or territories such as Singapore, Thailand, Philippines, Taiwan, Japan, South Korea, Iran, Italy, Spain, France, Germany, United Kingdom, United States of America, Australia, and Egypt, the sooner the proper actions were taken against the spread of the coronavirus, the smaller the size of the COVID-19 epidemic and its damage would likely be. Since the coronavirus is highly transmittable by droplets, more stringent control measures such as fast and mass testing, rigorous contact tracing, large-scale isolations, mandatory quarantines, travel restrictions or bans, border closing, social distancing, school closings, stay-at-home orders, and long-term lockdowns may be needed to contain the epidemic ultimately. Almost the whole world is now in the battle against the coronavirus as the global numbers of confirmed COVID-19 cases and deaths continue to rise speedily.

In such a disaster, various sectors of society should collaborate immediately to tackle the problem under the guidance of the government. Scientists and technology experts may try all the available tools to help the government fight the epidemic. The *basic reproduction number, R*_0_, for an infectious disease is the expected number of new cases infected directly from the index case in a susceptible population. As a measure of transmissibility, it tells us how quickly an infectious disease spreads out in various stages of the epidemic.^1^ Most importantly, *R*_0_ can be used to assess the effectiveness of the implemented control measures and to explore the possibility of pathogen mutations in an epidemic. When *R*_0_ < 1.0 persistently, the epidemic would be damped down soon.^1^ In essence, *R*_0_ is the driving force behind the epidemic curve. Its value usually varies over time during an epidemic. Hence, as biostatisticians, we started this investigation on January 27, 2020 with the aim to estimate concurrently the time-varying reproduction number, *R*_0_(*t*), over time during the ongoing epidemic of COVID-19 in China. We also initiated a similar research project for estimating concurrently the time-varying reproduction number, *R*_0_(*t*), over time during the ongoing epidemic of COVID-19 in 12 heavily-attacked countries outside China, which will be reported later.

## Methods

### Epidemic data

To collect all individual patient data, including personal contact history, during an epidemic is a tough and crucial task. As revealed in the report of the Novel Coronavirus Pneumonia Emergency Response Epidemiology Team, great efforts have previously been made in China to build up China’s Infectious Disease Information System, which is very helpful in the management of nationwide patient data during this epidemic of a large size.^2^ Such highly confidential and miscellaneous data are not made available for unauthorized researchers during the COVID-19 epidemic. Nevertheless, since January 19, 2020, the numbers of new confirmed cases, total confirmed cases, new suspected cases, total suspected cases, new deaths, total deaths, new recoveries, and total recoveries have been announced daily by the Chinese Center for Disease Control and Prevention (China CDC) on its website of “Tracking the Epidemic” (http://weekly.chinacdc.cn/news/TrackingtheEpidemic.htm). Thus, as listed in Supplementary Appendix 1, we extracted such publicly available data of this ongoing epidemic from the above website for the duration of January 19, 2020 and March 14, 2020 in this study. Yet, we had to make some minor corrections, marked with red asterisks, to reconcile the daily new counts with the daily total counts numerically.

### Epidemic analysis

Statistical analysis was performed using the R 3.6.3 software (R Foundation for Statistical Computing, Vienna, Austria). The distributional properties of counts were presented by the mean, standard deviation (SD), minimum, the first quartile (Q1), median, the third quartile (Q3), and maximum. Instead of developing any advanced methods specific to this epidemic, we tried to find an available easy-to-use tool to monitor the progress of the ongoing COVID-19 epidemic as soon as possible. As listed on the Comprehensive R Archive Network (CRAN) (https://cran.r-project.org/), several R packages might be used to compute basic reproduction numbers of an epidemic in R, including argo, epibasix, EpiCurve, EpiEstim, EpiILM, EpiILMCT, epimdr,^3^ epinet, epiR, EpiReport, epitools, epitrix, incidence, mem, memapp, *R*_0_, and surveillance. We chose the incidence (version 1.7.0) and EpiEstim (version 2.2-1) packages to estimate *R*_0_*(t)* in R during the ongoing COVID-19 epidemic in China due to their methodological soundness and computational simplicity for rapid analysis.^4,5^ Our R code was listed in Supplementary Appendix 2 for check and re-uses.

First, we laid out the conceptual framework below. In an epidemic of infectious disease, any susceptible subject who becomes a patient usually goes through the following three stages: infection, development of symptoms, and diagnosis of the disease. Theoretically, to estimate *R*_0_ or *R*_0_(*t*), we need the information about the distribution of *generation time* (GT), which is the time interval between the infection of the index case and the infection of the next case infected directly from the index case.^5^ Yet, the time of infection is most likely unavailable or inaccurate, and thus investigators collect the data about the distribution of *serial interval* (SI) instead, which is the time interval between the symptom onset of the index case and the symptom onset of the next case infected directly from the index case.^5^ Nevertheless, the data of symptom onset are not publically available and almost always have the problem of delayed reporting in any ongoing epidemic of infectious disease because they are usually recorded at diagnosis.^2^ Hence, we took a common approach in statistics to tackle this problem by specifying the best plausible distributions of SI according to the results obtained from previous studies of similar epidemics, and then applied the novel estimation method implemented in the EpiEstim package to the data of daily new confirmed cases in practice.^4,5^

Next, we considered two plausible scenarios for studying the ongoing COVID-19 epidemic in China. The estimate_R function of the EpiEstim package assumes a Gamma distribution for SI by default to approximate the infectivity profile.^4^ Technically, the transmission of an infectious disease is modeled with a Poisson process.^5^ When we choose a Gamma prior distribution for SI, the Bayesian statistical inference leads to a simple analytical expression for the Gamma posterior distribution of *R*_0_(*t*).^5^ In the first scenario, we specified the mean (SD) of the Gamma distribution for SI to be 8.4 (3.8) days to mimic the 2003 epidemic of the severe acute respiratory syndrome (SARS) in Hong Kong.^5^ Then, in the second scenario, we specified the mean (SD) of the Gamma distribution for SI to be 2.6 (1.5) days to mimic the 1918 pandemic of influenza in Baltimore, Maryland.^5^ Given an observed series of daily new confirmed cases, the shorter SI, the smaller *R*_0_(*t*). According to the current understanding, the transmissibility of COVID-19 was higher than SARS but lower than influenza.^1^ Hence, although we did not know the true distribution(s) of SI for the ongoing epidemic of COVID-19 in China,^6^ these two scenarios helped us catch the behavior pattern of this epidemic along with the time evolution.

## Results

### Epidemic data

We subtracted one day from the reporting dates in the COVID-19 epidemic data of China CDC to obtain the “dates” on which the daily new confirmed cases actually occurred. In Table 1, we listed the sample statistics of the daily new confirmed cases, total confirmed cases, new suspected cases, total suspected cases, new deaths, total deaths, new recoveries, and total recoveries during January 19-March 14 from the epidemic data of COVID-19 in China. Those statistics revealed the magnitude and impact of the ongoing epidemic over those 56 days. Yet, the clinically diagnosed cases (Hubei Province only) were added to the daily counts of new confirmed cases during February 12–16, 2020.^2^ Thus, the maximum value of new confirmed cases went up dramatically to 15,151 on February 12, 2020. All the counts of total confirmed cases, total deaths, and total recoveries were monotonically increasing, except the count of total suspected cases.

**Table 1.** Descriptive statistics for the epidemic data of the coronavirus disease 2019 (COVID-19) in China from January 19, 2020 to March 14, 2020 (*n* = 56).

| **Daily Counts\*** | **Mean** | **SD** | **Minimum** | **Q1** | **Median** | **Q3** | **Maximum** |
| --- | --- | --- | --- | --- | --- | --- | --- |
| New Confirmed Cases | 1446.91 | 2229.39 | 8 | 142.00 | 610.50 | 2060.75 | 15151* |
| Total Confirmed Cases | 50776.25 | 32313.72 | 214 | 16498.75 | 69524.00 | 80069.50 | 81027* |
| New Suspected Cases | 1736.91 | 1809.64 | 0 | 131.25 | 1151.50 | 3390.50 | 5328 |
| Total Suspected Cases | 7686.79 | 8927.01 | 0 | 515.50 | 3791.00 | 12484.00 | 28942 |
| New Deaths | 57.13 | 43.67 | 0 | 23.50 | 43.50 | 97.00 | 150 |
| Total Deaths | 1633.73 | 1226.35 | 6 | 346.75 | 1717.50 | 2880.50 | 3199 |
| New Recoveries | 1194.84 | 1011.94 | 0 | 131.50 | 1318.00 | 1829.50 | 3622 |
| Total Recoveries | 21405.84 | 23650.20 | 25 | 438.25 | 10131.50 | 42334.25 | 66911 |

Abbreviation: Q1 = the first quartile, Q3 = the third quartile, and SD = standard deviation.
\* The clinically diagnosed cases (Hubei Province only) were added to the daily counts of new confirmed cases from February 12 to February 16, 2020.

### Epidemic analysis

In Figure 1, the epidemic curve of daily new confirmed cases peaked around February 4–5, 2020. Then, it declined gradually, except for the very high peak on February 12, 2020 (in the dark red color) due to the added clinically diagnosed cases of the Hubei Province.

**Figure 1.**
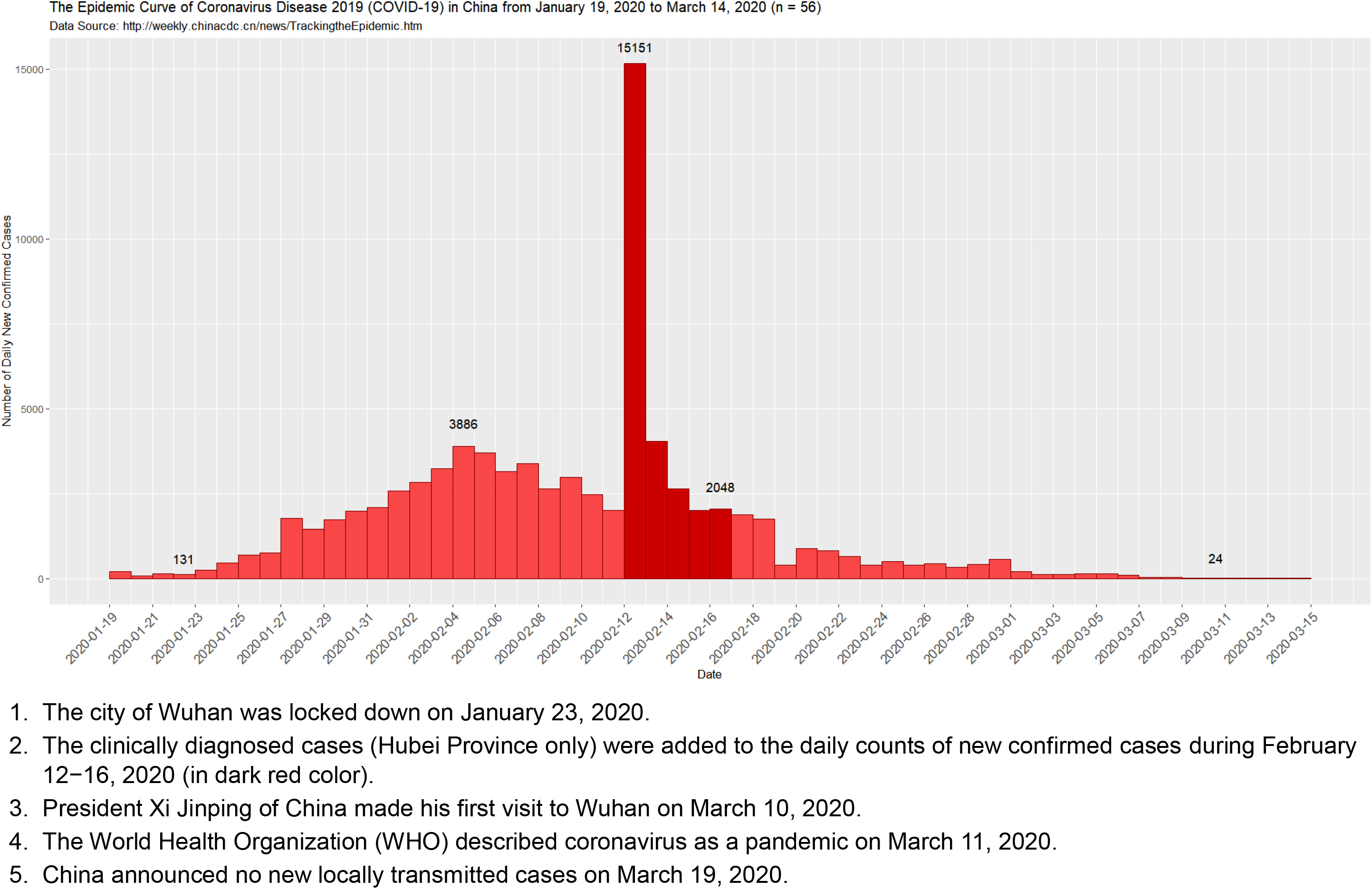
The epidemic curve of the coronavirus disease 2019 (COVID-19) in China from January 19, 2020 to March 14, 2020.

Next, under the above specified two scenarios, we plotted the daily estimates of the time-varying reproduction numbers, *R*_0_(t), over sliding weekly windows,^4,5^ from January 19 to March 14, 2020 in the middle panels of Figures 2A and 2B for the ongoing COVID-19 epidemic in China. Namely, at any given day of the epidemic curve, *R*_0_*(t)* was estimated for the weekly window ending on that day.^4,5^ The estimated *R*_0_*(t)* was not shown from the very beginning of the epidemic because precise estimation was not possible in that period.^5^ Specifically, the blue lines showed the posterior means of *R*_0_(*t*), the grey zones represented the 95% credible intervals (CrI), and the black horizontal dashed lines indicated the threshold value of *R*_0_ = 1.0.^4,5^ In addition, the first and third panels of Figures 2A and 2B were the miniaturized epidemic curve of daily new confirmed cases (see Figure 1) and the distributions of SI used for the estimation of *R*_0_(*t*).^4,5^ Intriguingly, both curves of the estimated R_0_(t) went down to be less than 1.0 around February 17–18, 2020.

**Figure 2.**
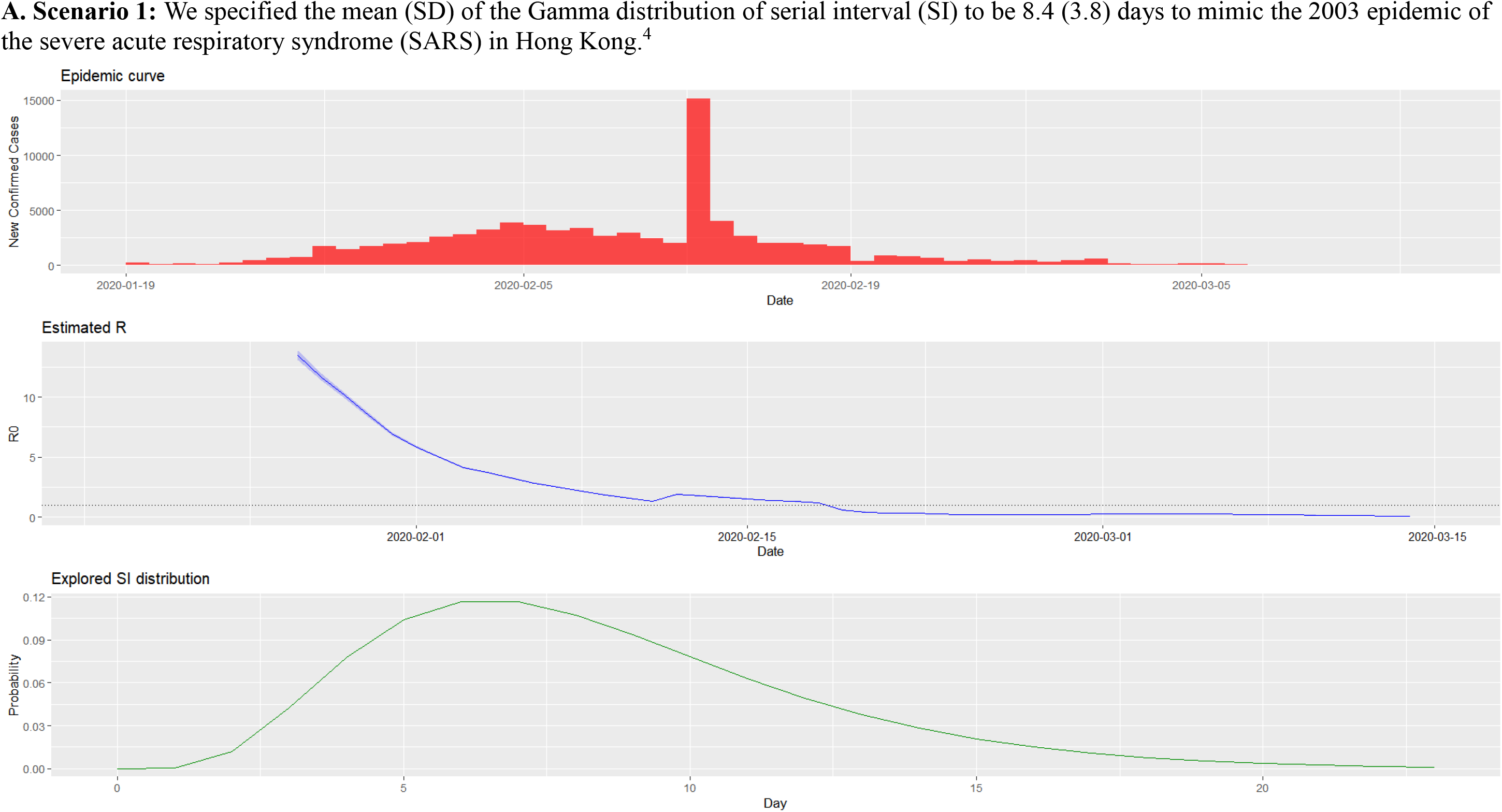
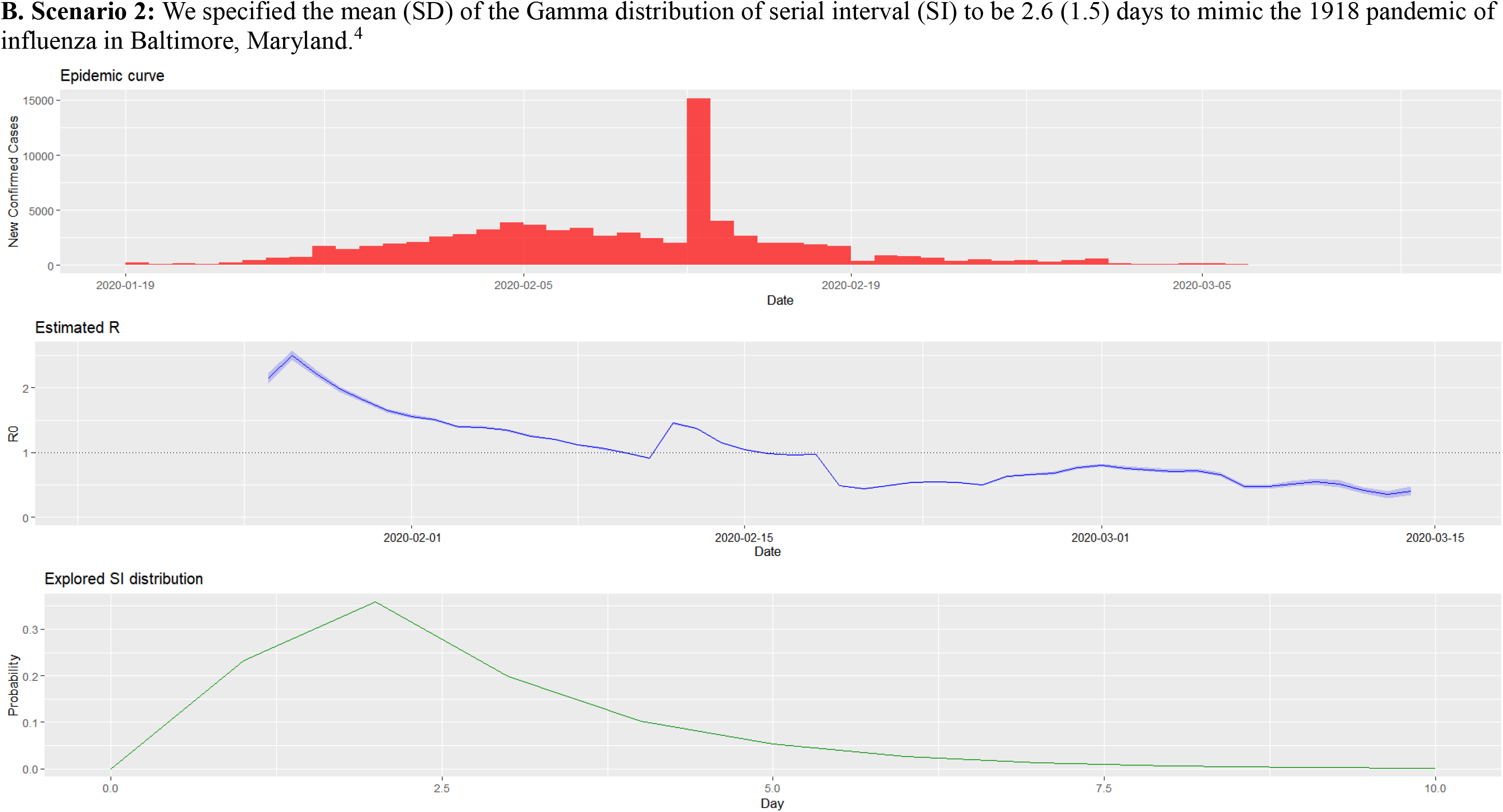
The estimated time-varying reproduction number during the ongoing epidemic of the coronavirus disease 2019 (COVID-19) in China from January 19, 2020 to March 14, 2020 under two scenarios.

Finally, in Tables 2A and 2B, we displayed the estimated parameters of the Gamma posterior distributions of *R*_0_(*t*), over sliding weekly windows, from January 19 to March 14, 2020 under those two specified scenarios for scrutinization.^4,5^

**Table 2.**
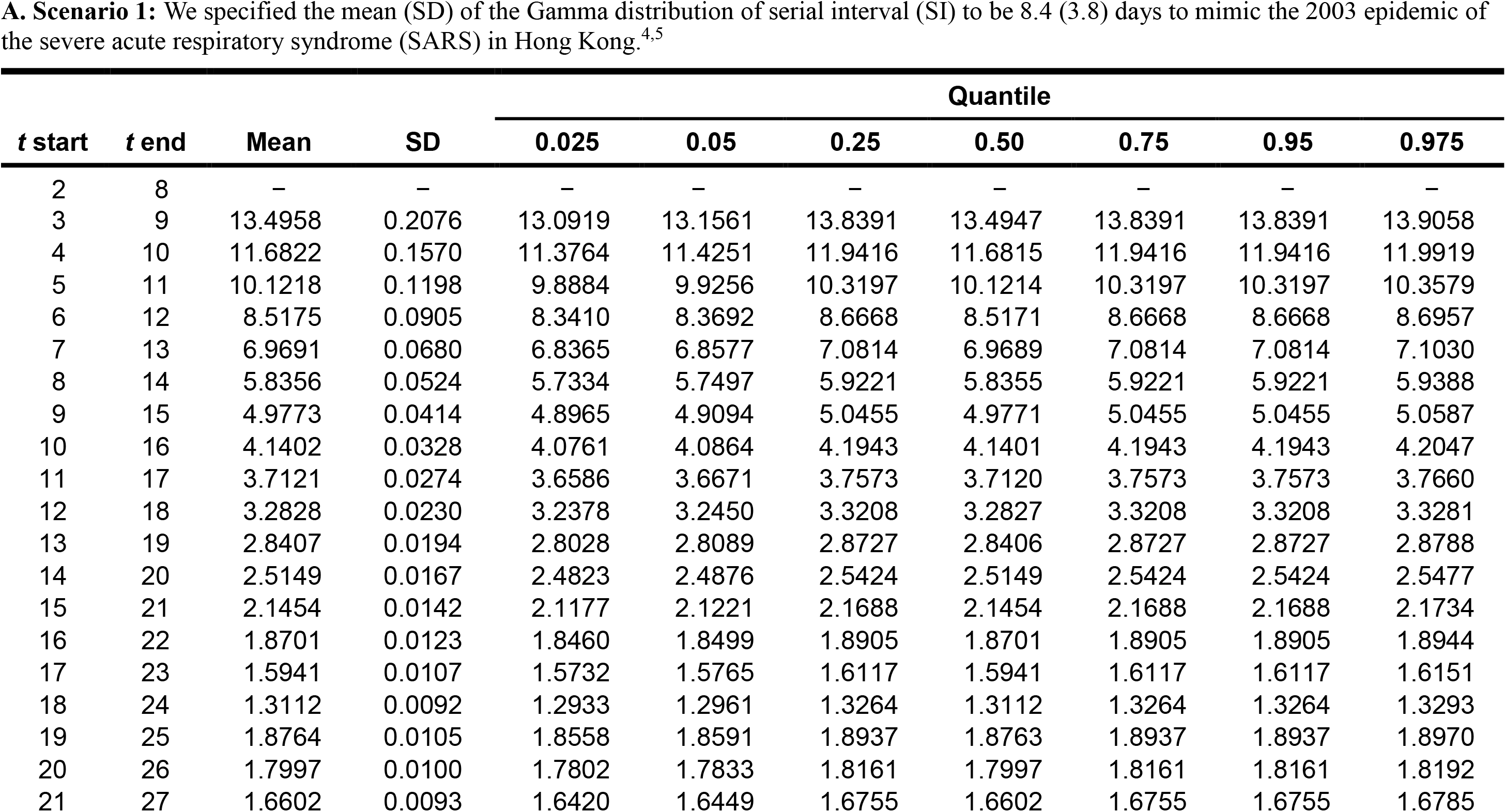
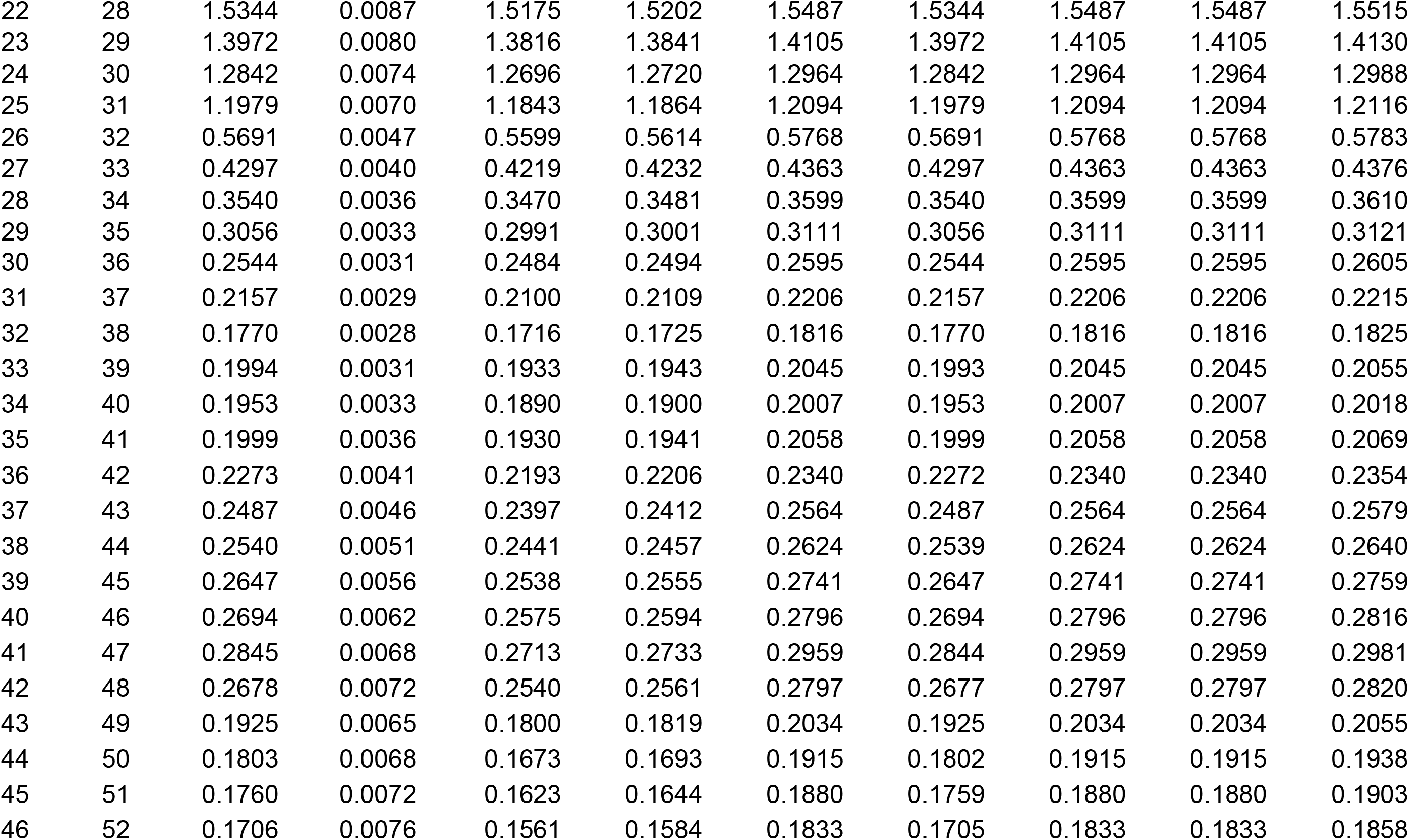
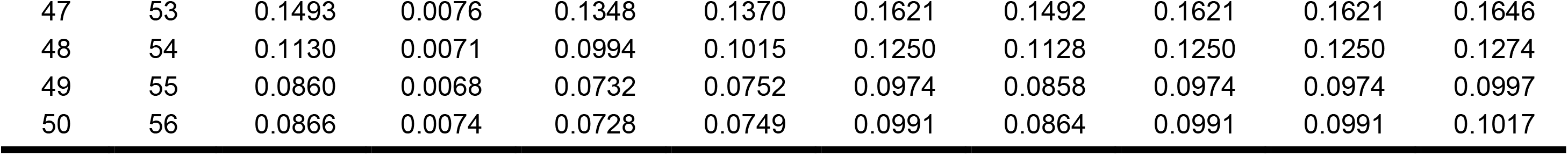
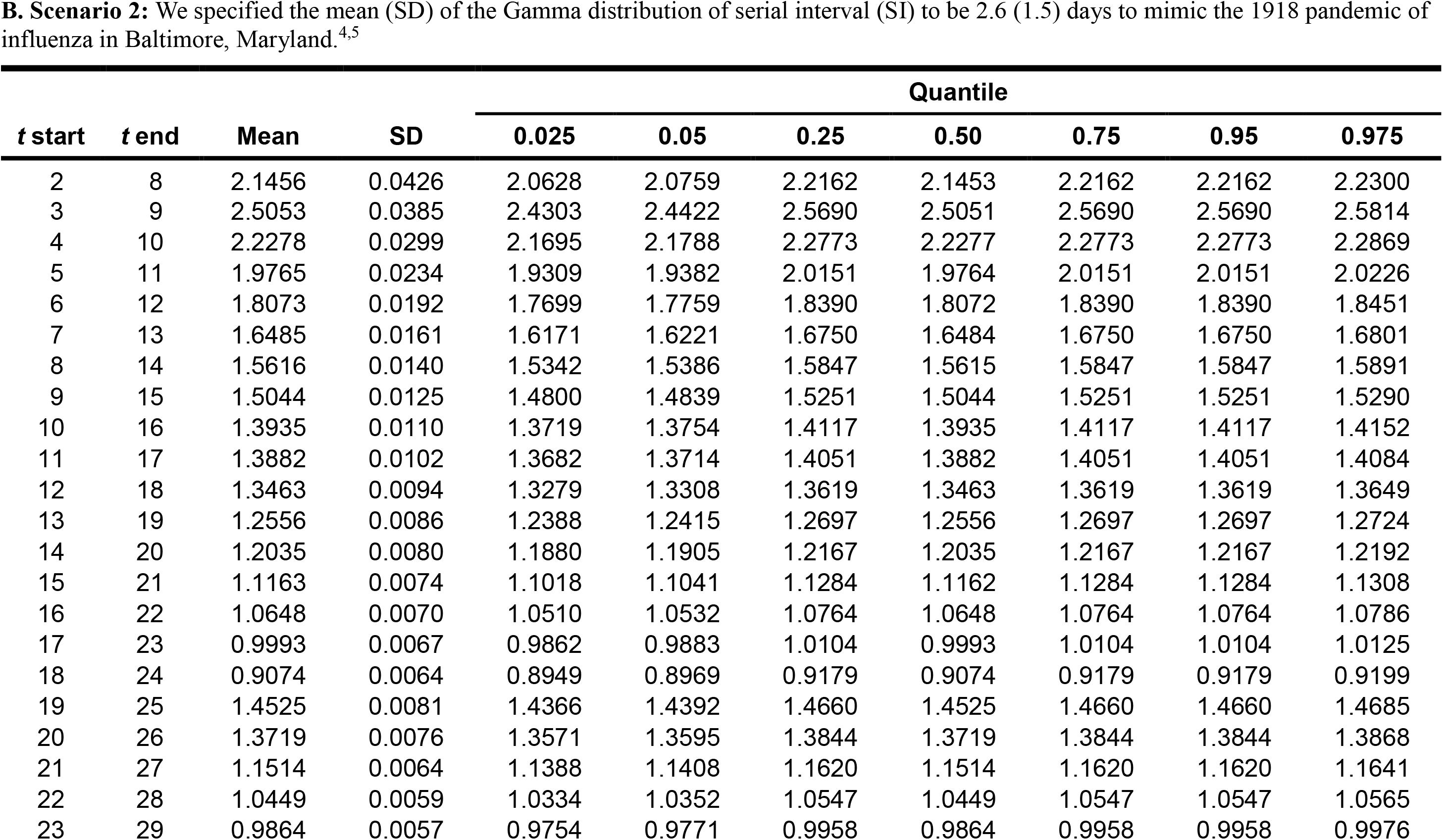
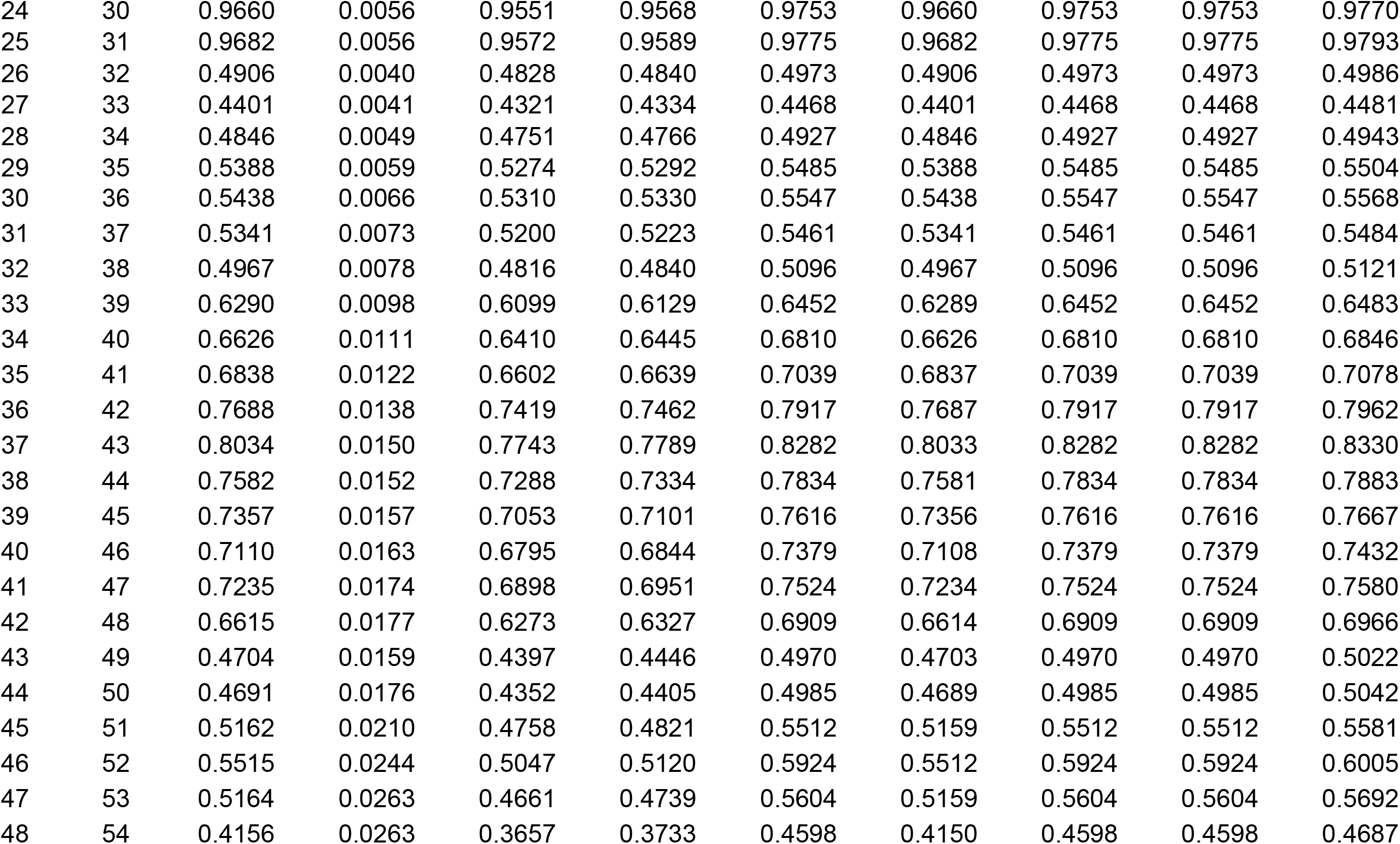
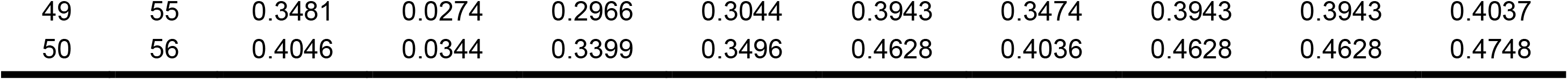
The estimated distributional parameters of the time-varying reproduction number during the ongoing epidemic of the coronavirus disease 2019 (COVID-19) in China from January 19, 2020 to March 14, 2020 under two scenarios.

| *t* start | *t* end | Mean | SD | Quantile |  |  |  |  |  |  |
| --- | --- | --- | --- | --- | --- | --- | --- | --- | --- | --- |
|  |  |  |  | 0.025 | 0.05 | 0.25 | 0.50 | 0.75 | 0.95 | 0.975 |
| 2 | 8 | – | – | – | – | – | – | – | – | – |
| 3 | 9 | 13.4958 | 0.2076 | 13.0919 | 13.1561 | 13.8391 | 13.4947 | 13.8391 | 13.8391 | 13.9058 |
| 4 | 10 | 11.6822 | 0.1570 | 11.3764 | 11.4251 | 11.9416 | 11.6815 | 11.9416 | 11.9416 | 11.9919 |
| 5 | 11 | 10.1218 | 0.1198 | 9.8884 | 9.9256 | 10.3197 | 10.1214 | 10.3197 | 10.3197 | 10.3579 |
| 6 | 12 | 8.5175 | 0.0905 | 8.3410 | 8.3692 | 8.6668 | 8.5171 | 8.6668 | 8.6668 | 8.6957 |
| 7 | 13 | 6.9691 | 0.0680 | 6.8365 | 6.8577 | 7.0814 | 6.9689 | 7.0814 | 7.0814 | 7.1030 |
| 8 | 14 | 5.8356 | 0.0524 | 5.7334 | 5.7497 | 5.9221 | 5.8355 | 5.9221 | 5.9221 | 5.9388 |
| 9 | 15 | 4.9773 | 0.0414 | 4.8965 | 4.9094 | 5.0455 | 4.9771 | 5.0455 | 5.0455 | 5.0587 |
| 10 | 16 | 4.1402 | 0.0328 | 4.0761 | 4.0864 | 4.1943 | 4.1401 | 4.1943 | 4.1943 | 4.2047 |
| 11 | 17 | 3.7121 | 0.0274 | 3.6586 | 3.6671 | 3.7573 | 3.7120 | 3.7573 | 3.7573 | 3.7660 |
| 12 | 18 | 3.2828 | 0.0230 | 3.2378 | 3.2450 | 3.3208 | 3.2827 | 3.3208 | 3.3208 | 3.3281 |
| 13 | 19 | 2.8407 | 0.0194 | 2.8028 | 2.8089 | 2.8727 | 2.8406 | 2.8727 | 2.8727 | 2.8788 |
| 14 | 20 | 2.5149 | 0.0167 | 2.4823 | 2.4876 | 2.5424 | 2.5149 | 2.5424 | 2.5424 | 2.5477 |
| 15 | 21 | 2.1454 | 0.0142 | 2.1177 | 2.1221 | 2.1688 | 2.1454 | 2.1688 | 2.1688 | 2.1734 |
| 16 | 22 | 1.8701 | 0.0123 | 1.8460 | 1.8499 | 1.8905 | 1.8701 | 1.8905 | 1.8905 | 1.8944 |
| 17 | 23 | 1.5941 | 0.0107 | 1.5732 | 1.5765 | 1.6117 | 1.5941 | 1.6117 | 1.6117 | 1.6151 |
| 18 | 24 | 1.3112 | 0.0092 | 1.2933 | 1.2961 | 1.3264 | 1.3112 | 1.3264 | 1.3264 | 1.3293 |
| 19 | 25 | 1.8764 | 0.0105 | 1.8558 | 1.8591 | 1.8937 | 1.8763 | 1.8937 | 1.8937 | 1.8970 |
| 20 | 26 | 1.7997 | 0.0100 | 1.7802 | 1.7833 | 1.8161 | 1.7997 | 1.8161 | 1.8161 | 1.8192 |
| 21 | 27 | 1.6602 | 0.0093 | 1.6420 | 1.6449 | 1.6755 | 1.6602 | 1.6755 | 1.6755 | 1.6785 |
| 22 | 28 | 1.5344 | 0.0087 | 1.5175 | 1.5202 | 1.5487 | 1.5344 | 1.5487 | 1.5487 | 1.5515 |
| 23 | 29 | 1.3972 | 0.0080 | 1.3816 | 1.3841 | 1.4105 | 1.3972 | 1.4105 | 1.4105 | 1.4130 |
| 24 | 30 | 1.2842 | 0.0074 | 1.2696 | 1.2720 | 1.2964 | 1.2842 | 1.2964 | 1.2964 | 1.2988 |
| 25 | 31 | 1.1979 | 0.0070 | 1.1843 | 1.1864 | 1.2094 | 1.1979 | 1.2094 | 1.2094 | 1.2116 |
| 26 | 32 | 0.5691 | 0.0047 | 0.5599 | 0.5614 | 0.5768 | 0.5691 | 0.5768 | 0.5768 | 0.5783 |
| 27 | 33 | 0.4297 | 0.0040 | 0.4219 | 0.4232 | 0.4363 | 0.4297 | 0.4363 | 0.4363 | 0.4376 |
| 28 | 34 | 0.3540 | 0.0036 | 0.3470 | 0.3481 | 0.3599 | 0.3540 | 0.3599 | 0.3599 | 0.3610 |
| 29 | 35 | 0.3056 | 0.0033 | 0.2991 | 0.3001 | 0.3111 | 0.3056 | 0.3111 | 0.3111 | 0.3121 |
| 30 | 36 | 0.2544 | 0.0031 | 0.2484 | 0.2494 | 0.2595 | 0.2544 | 0.2595 | 0.2595 | 0.2605 |
| 31 | 37 | 0.2157 | 0.0029 | 0.2100 | 0.2109 | 0.2206 | 0.2157 | 0.2206 | 0.2206 | 0.2215 |
| 32 | 38 | 0.1770 | 0.0028 | 0.1716 | 0.1725 | 0.1816 | 0.1770 | 0.1816 | 0.1816 | 0.1825 |
| 33 | 39 | 0.1994 | 0.0031 | 0.1933 | 0.1943 | 0.2045 | 0.1993 | 0.2045 | 0.2045 | 0.2055 |
| 34 | 40 | 0.1953 | 0.0033 | 0.1890 | 0.1900 | 0.2007 | 0.1953 | 0.2007 | 0.2007 | 0.2018 |
| 35 | 41 | 0.1999 | 0.0036 | 0.1930 | 0.1941 | 0.2058 | 0.1999 | 0.2058 | 0.2058 | 0.2069 |
| 36 | 42 | 0.2273 | 0.0041 | 0.2193 | 0.2206 | 0.2340 | 0.2272 | 0.2340 | 0.2340 | 0.2354 |
| 37 | 43 | 0.2487 | 0.0046 | 0.2397 | 0.2412 | 0.2564 | 0.2487 | 0.2564 | 0.2564 | 0.2579 |
| 38 | 44 | 0.2540 | 0.0051 | 0.2441 | 0.2457 | 0.2624 | 0.2539 | 0.2624 | 0.2624 | 0.2640 |
| 39 | 45 | 0.2647 | 0.0056 | 0.2538 | 0.2555 | 0.2741 | 0.2647 | 0.2741 | 0.2741 | 0.2759 |
| 40 | 46 | 0.2694 | 0.0062 | 0.2575 | 0.2594 | 0.2796 | 0.2694 | 0.2796 | 0.2796 | 0.2816 |
| 41 | 47 | 0.2845 | 0.0068 | 0.2713 | 0.2733 | 0.2959 | 0.2844 | 0.2959 | 0.2959 | 0.2981 |
| 42 | 48 | 0.2678 | 0.0072 | 0.2540 | 0.2561 | 0.2797 | 0.2677 | 0.2797 | 0.2797 | 0.2820 |
| 43 | 49 | 0.1925 | 0.0065 | 0.1800 | 0.1819 | 0.2034 | 0.1925 | 0.2034 | 0.2034 | 0.2055 |
| 44 | 50 | 0.1803 | 0.0068 | 0.1673 | 0.1693 | 0.1915 | 0.1802 | 0.1915 | 0.1915 | 0.1938 |
| 45 | 51 | 0.1760 | 0.0072 | 0.1623 | 0.1644 | 0.1880 | 0.1759 | 0.1880 | 0.1880 | 0.1903 |
| 46 | 52 | 0.1706 | 0.0076 | 0.1561 | 0.1584 | 0.1833 | 0.1705 | 0.1833 | 0.1833 | 0.1858 |
| 47 | 53 | 0.1493 | 0.0076 | 0.1348 | 0.1370 | 0.1621 | 0.1492 | 0.1621 | 0.1621 | 0.1646 |
| 48 | 54 | 0.1130 | 0.0071 | 0.0994 | 0.1015 | 0.1250 | 0.1128 | 0.1250 | 0.1250 | 0.1274 |
| 49 | 55 | 0.0860 | 0.0068 | 0.0732 | 0.0752 | 0.0974 | 0.0858 | 0.0974 | 0.0974 | 0.0997 |
| 50 | 56 | 0.0866 | 0.0074 | 0.0728 | 0.0749 | 0.0991 | 0.0864 | 0.0991 | 0.0991 | 0.1017 |
|  |  |  |  | 0.025 | 0.05 | 0.25 | 0.50 | 0.75 | 0.95 | 0.975 |
| 2 | 8 | 2.1456 | 0.0426 | 2.0628 | 2.0759 | 2.2162 | 2.1453 | 2.2162 | 2.2162 | 2.2300 |
| 3 | 9 | 2.5053 | 0.0385 | 2.4303 | 2.4422 | 2.5690 | 2.5051 | 2.5690 | 2.5690 | 2.5814 |
| 4 | 10 | 2.2278 | 0.0299 | 2.1695 | 2.1788 | 2.2773 | 2.2277 | 2.2773 | 2.2773 | 2.2869 |
| 5 | 11 | 1.9765 | 0.0234 | 1.9309 | 1.9382 | 2.0151 | 1.9764 | 2.0151 | 2.0151 | 2.0226 |
| 6 | 12 | 1.8073 | 0.0192 | 1.7699 | 1.7759 | 1.8390 | 1.8072 | 1.8390 | 1.8390 | 1.8451 |
| 7 | 13 | 1.6485 | 0.0161 | 1.6171 | 1.6221 | 1.6750 | 1.6484 | 1.6750 | 1.6750 | 1.6801 |
| 8 | 14 | 1.5616 | 0.0140 | 1.5342 | 1.5386 | 1.5847 | 1.5615 | 1.5847 | 1.5847 | 1.5891 |
| 9 | 15 | 1.5044 | 0.0125 | 1.4800 | 1.4839 | 1.5251 | 1.5044 | 1.5251 | 1.5251 | 1.5290 |
| 10 | 16 | 1.3935 | 0.0110 | 1.3719 | 1.3754 | 1.4117 | 1.3935 | 1.4117 | 1.4117 | 1.4152 |
| 11 | 17 | 1.3882 | 0.0102 | 1.3682 | 1.3714 | 1.4051 | 1.3882 | 1.4051 | 1.4051 | 1.4084 |
| 12 | 18 | 1.3463 | 0.0094 | 1.3279 | 1.3308 | 1.3619 | 1.3463 | 1.3619 | 1.3619 | 1.3649 |
| 13 | 19 | 1.2556 | 0.0086 | 1.2388 | 1.2415 | 1.2697 | 1.2556 | 1.2697 | 1.2697 | 1.2724 |
| 14 | 20 | 1.2035 | 0.0080 | 1.1880 | 1.1905 | 1.2167 | 1.2035 | 1.2167 | 1.2167 | 1.2192 |
| 15 | 21 | 1.1163 | 0.0074 | 1.1018 | 1.1041 | 1.1284 | 1.1162 | 1.1284 | 1.1284 | 1.1308 |
| 16 | 22 | 1.0648 | 0.0070 | 1.0510 | 1.0532 | 1.0764 | 1.0648 | 1.0764 | 1.0764 | 1.0786 |
| 17 | 23 | 0.9993 | 0.0067 | 0.9862 | 0.9883 | 1.0104 | 0.9993 | 1.0104 | 1.0104 | 1.0125 |
| 18 | 24 | 0.9074 | 0.0064 | 0.8949 | 0.8969 | 0.9179 | 0.9074 | 0.9179 | 0.9179 | 0.9199 |
| 19 | 25 | 1.4525 | 0.0081 | 1.4366 | 1.4392 | 1.4660 | 1.4525 | 1.4660 | 1.4660 | 1.4685 |
| 20 | 26 | 1.3719 | 0.0076 | 1.3571 | 1.3595 | 1.3844 | 1.3719 | 1.3844 | 1.3844 | 1.3868 |
| 21 | 27 | 1.1514 | 0.0064 | 1.1388 | 1.1408 | 1.1620 | 1.1514 | 1.1620 | 1.1620 | 1.1641 |
| 22 | 28 | 1.0449 | 0.0059 | 1.0334 | 1.0352 | 1.0547 | 1.0449 | 1.0547 | 1.0547 | 1.0565 |
| 23 | 29 | 0.9864 | 0.0057 | 0.9754 | 0.9771 | 0.9958 | 0.9864 | 0.9958 | 0.9958 | 0.9976 |
| 24 | 30 | 0.9660 | 0.0056 | 0.9551 | 0.9568 | 0.9753 | 0.9660 | 0.9753 | 0.9753 | 0.9770 |
| 25 | 31 | 0.9682 | 0.0056 | 0.9572 | 0.9589 | 0.9775 | 0.9682 | 0.9775 | 0.9775 | 0.9793 |
| 26 | 32 | 0.4906 | 0.0040 | 0.4828 | 0.4840 | 0.4973 | 0.4906 | 0.4973 | 0.4973 | 0.4986 |
| 27 | 33 | 0.4401 | 0.0041 | 0.4321 | 0.4334 | 0.4468 | 0.4401 | 0.4468 | 0.4468 | 0.4481 |
| 28 | 34 | 0.4846 | 0.0049 | 0.4751 | 0.4766 | 0.4927 | 0.4846 | 0.4927 | 0.4927 | 0.4943 |
| 29 | 35 | 0.5388 | 0.0059 | 0.5274 | 0.5292 | 0.5485 | 0.5388 | 0.5485 | 0.5485 | 0.5504 |
| 30 | 36 | 0.5438 | 0.0066 | 0.5310 | 0.5330 | 0.5547 | 0.5438 | 0.5547 | 0.5547 | 0.5568 |
| 31 | 37 | 0.5341 | 0.0073 | 0.5200 | 0.5223 | 0.5461 | 0.5341 | 0.5461 | 0.5461 | 0.5484 |
| 32 | 38 | 0.4967 | 0.0078 | 0.4816 | 0.4840 | 0.5096 | 0.4967 | 0.5096 | 0.5096 | 0.5121 |
| 33 | 39 | 0.6290 | 0.0098 | 0.6099 | 0.6129 | 0.6452 | 0.6289 | 0.6452 | 0.6452 | 0.6483 |
| 34 | 40 | 0.6626 | 0.0111 | 0.6410 | 0.6445 | 0.6810 | 0.6626 | 0.6810 | 0.6810 | 0.6846 |
| 35 | 41 | 0.6838 | 0.0122 | 0.6602 | 0.6639 | 0.7039 | 0.6837 | 0.7039 | 0.7039 | 0.7078 |
| 36 | 42 | 0.7688 | 0.0138 | 0.7419 | 0.7462 | 0.7917 | 0.7687 | 0.7917 | 0.7917 | 0.7962 |
| 37 | 43 | 0.8034 | 0.0150 | 0.7743 | 0.7789 | 0.8282 | 0.8033 | 0.8282 | 0.8282 | 0.8330 |
| 38 | 44 | 0.7582 | 0.0152 | 0.7288 | 0.7334 | 0.7834 | 0.7581 | 0.7834 | 0.7834 | 0.7883 |
| 39 | 45 | 0.7357 | 0.0157 | 0.7053 | 0.7101 | 0.7616 | 0.7356 | 0.7616 | 0.7616 | 0.7667 |
| 40 | 46 | 0.7110 | 0.0163 | 0.6795 | 0.6844 | 0.7379 | 0.7108 | 0.7379 | 0.7379 | 0.7432 |
| 41 | 47 | 0.7235 | 0.0174 | 0.6898 | 0.6951 | 0.7524 | 0.7234 | 0.7524 | 0.7524 | 0.7580 |
| 42 | 48 | 0.6615 | 0.0177 | 0.6273 | 0.6327 | 0.6909 | 0.6614 | 0.6909 | 0.6909 | 0.6966 |
| 43 | 49 | 0.4704 | 0.0159 | 0.4397 | 0.4446 | 0.4970 | 0.4703 | 0.4970 | 0.4970 | 0.5022 |
| 44 | 50 | 0.4691 | 0.0176 | 0.4352 | 0.4405 | 0.4985 | 0.4689 | 0.4985 | 0.4985 | 0.5042 |
| 45 | 51 | 0.5162 | 0.0210 | 0.4758 | 0.4821 | 0.5512 | 0.5159 | 0.5512 | 0.5512 | 0.5581 |
| 46 | 52 | 0.5515 | 0.0244 | 0.5047 | 0.5120 | 0.5924 | 0.5512 | 0.5924 | 0.5924 | 0.6005 |
| 47 | 53 | 0.5164 | 0.0263 | 0.4661 | 0.4739 | 0.5604 | 0.5159 | 0.5604 | 0.5604 | 0.5692 |
| 48 | 54 | 0.4156 | 0.0263 | 0.3657 | 0.3733 | 0.4598 | 0.4150 | 0.4598 | 0.4598 | 0.4687 |
| 49 | 55 | 0.3481 | 0.0274 | 0.2966 | 0.3044 | 0.3943 | 0.3474 | 0.3943 | 0.3943 | 0.4037 |
| 50 | 56 | 0.4046 | 0.0344 | 0.3399 | 0.3496 | 0.4628 | 0.4036 | 0.4628 | 0.4628 | 0.4748 |

## Discussion

Almost everyone was susceptible to the novel COVID-19 and this was one of the reasons why the COVID-19 epidemic occurred in many places and caused public panics in China and worldwide. In terms of population size, the depletion due to death or recovery might be negligible in China. And, there were no imported cases of COVID-19 in China. These features made the task of modeling this epidemic in China relatively easier. However, since the COVID-19 was new to human society, its diagnostic criteria, control measures, and medical cares were inevitably changing during the epidemic as the knowledge and experience about it were accumulated continuously.^2^ We decided not to exclude the added clinically diagnosed cases of the Hubei Province during February 12-16, 2020 for the robustness of our epidemic analysis. Besides, many recoveries were probably not susceptible any more in the late phase of the epidemic. Thus, the result obtained in this study was merely a rough estimate of *R*_0_(*t*). Nevertheless, our findings (esp., Figure 2B) were consistent with the earlier estimates of *R*_0_ for the COVID-19 epidemic in China such as 2.24 (95% confidence interval [CI]: 1.96–2.55)–3.58 (95% CI: 2.89–4.39),^6^ 2.2 (95% CI: 1.4–3.9),^7^ 2.6 (uncertainty range: 1.5–3.5),^8^ 2.68 (95% CrI: 2.47−2.86),^9^ 1.4–2.5 (WHO),^10^ 2.0–3.3,^10^ 2.2 (90% CI: 1.4–3.8),^10^ 6.47 (95% CI: 5.71–7.23),^10^ 3.11 (90% CI: 2.39–4.13),^10^ and 2.0.^10^

This study had several limitations because we relied on some assumptions to make a rapid analysis of this ongoing epidemic feasible. First, we assumed that all new cases of COVID-19 in China were detected and put into the counts of daily new confirmed cases correctly. However, asymptomatic or mild cases of COVID-19 were likely undetected, and thus under-reported, especially in the early phase of this epidemic.^6,11^ In some countries, a lack of diagnostic test kits for the SARS-CoV-2 and a shortage of qualified manpower for fast testing could also cause under-reporting or delay in reporting. Second, we admitted the time delay in our estimates of *R*_0_(*t*) for the COVID-19 epidemic in China due to the following two time lags: (1) the duration between the time of infection and the time of symptom onset (i.e., the *incubation period* of infection) if infectiousness began around the time of symptom onset^5^ and (2) the duration between the time of symptom onset and the time of diagnosis.^2^ Nevertheless, if asymptomatic carriers could transmit the COVID-19,^12^ the first time lag would be shorter and it became the duration between the time of infection and the time of becoming infectious (i.e., the *latent period* of infection). Moreover, the time interval from symptom onset to diagnosis was shorter and shorter due to the full alert of society and the faster diagnostic tests.^2^ Third, we assumed that the distribution of SI did not change considerably over time as the epidemic progressed. However, as for the SARS epidemic in Singapore, the SI tended to be shorter after control measures were implemented.^13^ The SI also depended on the amount of infecting dose, the level of host immunity, and the frequency of person-to-person contacts. To summarize, most of these limitations led our estimation of *R*_0_(*t*) into a more conservative context.

Looking back the epidemic curve in Figure 1, we could see that the lockdown of Wuhan on January 23, 2020 was a very smart, brave, and quick move even though the numbers of new confirmed cases and total confirmed cases were only 131 and 571 on January 22, 2020 in the very early phase of the COVID-19 epidemic in China. Then, adding clinically diagnosed cases of the Hubei Province to the daily counts of new confirmed cases during February 12–16, 2020 indicated that the capacity of medical care was strong enough at that time to help the suspected cases obtain proper medical care sooner by reducing the waiting time. As a result, the new confirmed cases had dropped sharply after February 18, 2020.

Next, as shown in Figures 2A and 2B, the estimated *R*_0_(*t*) began to drop just 2–4 days after the lockdown of Wuhan on January 23, 2020. Yet, the number of new confirmed cases was still climbing up day by day until it reached the peak around February 4–5, 2020. During this very painful time period, everyone was keen to know: When would the epidemic curve reach the hilltop?^14^ Although it was difficult to estimate the exact date when it would happen, the peak of 3,886 new confirmed cases on February 4, 2020 appeared right after the trend of the computed *R*_0_(*t*) was declining monotonically for about 10 days, and then the daily number of new confirmed cases began dropping, indicating that the COVID-19 epidemic had abated. Then, two weeks later, the estimated *R*_0_(*t*) surprisingly reduced to the level below 1.0 around February 17–18, 2020 in both scenarios. Our first epidemic analysis was done on February 24, 2020, which had led us to believe that the COVID-19 epidemic would end soon if the effective control measures were maintained and nothing else happened incidentally such as heavy case imports, super-spreaders, and virulent virus mutations. Finally, we observed that the COVID-19 epidemic in China closed up around March 7–8, 2020, indicating that the prompt and aggressive control measures of China^15-18^ were effective. In the end, China would win the battle against the coronavirus as long as its resurgence, if any, is well managed. In our opinion, seeing the estimated *R*_0_(*t*) going downhill is more informative than looking for the drops in the daily number of new confirmed cases during an ongoing epidemic of infectious disease. And, the consistent decline of the estimated *R*_0_(*t*) in trend over time is more important than the actual values of the estimated *R*_0_(*t*) themselves. The steeper the slope, the sooner the epidemic ends.

Although this study could not provide the most accurate results in a rigorous way, it sufficed for the pragmatic purpose from the public health viewpoint. We believed that it was an approximate answer to the right question. The estimate_R function of the EpiEstim package provided the option for including the data of imported cases in the estimation of *R*_0_(*t*).^4^ Refinements in the estimation of *R*_0_(*t*) can be made with the individual patient data, including personal contact history, whenever they are available for analysis.

Finally, we urged public health authorities and scientists worldwide to estimate time-varying reproduction numbers routinely during epidemics of infectious diseases and to report them daily on their websites such as the “Tracking the Epidemic” of China CDC for guiding the control strategies and reducing the unnecessary panic of the public until the end of the epidemic.^18^ Fitting complex transmission models of infectious disease dynamics to epidemic data with limited information about required parameters is a challenge.^3^ The results of such analyses may be difficult to generalize due to the context-specific assumptions made and it can be too slow to meet a pressing need during an epidemic.^5,19-22^ Thus, an easy-to-use tool for monitoring the COVID-19 epidemic is so important in practice.

Since the coronavirus has spread out globally, we should take the fresh lessons from China^15-22^, South Korea^23^, Italy^24^, and the United States of America^25,26^ and learn the experiences from the previous epidemics^1^ to mitigate its harm as much as possible. We may also use China as an example to anticipate the potential progression of the COVID-19 epidemic in a particular country. Control tactics and measures should be applied in line with local circumstances, but the same easy-to-use monitoring tool, *R*_0_(*t*), could be applied to many places. Let’s help each other to combat the COVID-19 pandemic together. After all, we are all in the same shaking boat now.

## Data Availability

The epidemic data were listed in Supplementary Appendix 1.

http://weekly.chinacdc.cn/news/TrackingtheEpidemic.htm

